# Conditional Generative Adversarial Networks for Individualized Treatment Effect Estimation and Treatment Selection

**DOI:** 10.1101/2020.09.28.20203075

**Authors:** Qiyang Ge, Xuelin Huang, Shenying Fang, Shicheng Guo, Yuanyuan Liu, Wei Lin, Momiao Xiong

## Abstract

Treatment response is heterogeneous. However the classical methods treat the treatment response as homogeneous and estimate the average treatment effects. The traditional methods are difficult to apply to precision oncology. The artificial intelligence (AI) is a powerful tool for precision oncology. It can accurately estimate the individualized treatment effects and learn optimal treatment choices. Therefore, the AI approach can substantially improve progress and treatment outcomes of patients. As one of AI approach, conditional generative adversarial nets for inference of individualized treatment effects (GANITE) have been developed. However, the GANITE can only deal with binary treatment and does not provide a tool for optimal treatment selection. To overcome these limitations, we modify conditional generative adversarial networks (MCGANs) to allow estimation of individualized effects of any types of treatments including binary, categorical and continuous treatments. We propose to use sparse techniques for selection of biomarkers that predict the best treatment for each patient. Simulations show that the CGANs outperform seven other state-of-the-art methods: linear regression (LR), Bayesian linear ridge regression (BLR), KNN, random forest classification (RF (C)), random forest regression (RF (R)), logistic regression (LogR) and support vector machine (SVM). To illustrate their applications, the proposed CGANs were applied to 256 patients with newly diagnosed acute myeloid leukemia (AML) who were treated with high dose ara-C (HDAC), Idarubicin (IDA) and both of these two treatments (HDAC+IDA) at M. D. Anderson Cancer Center. Our results showed that the MCGAN can more accurately and robustly estimate the individualized treatment effects than other state-of-the art methods. Several biomarkers such as GSK3, BILIRUBIN, SMAC are identified and a total of 30 biomarkers can explain 36.8% of treatment effect variation.

## Introduction

The traditional clinical management estimates the average treatment effects from observational data, assuming the complex disease is homogeneous (Hansen 2024; Kennedy et al. 2017; Liu et al. 2018; Luo and Zhu 2017; Rosenbaum and Rubin 1983; Diamond and Sekhon 2013). Alternative to traditional clinical management, “precision medicine” or “precision oncology” attempts to match the most accurate and effective treatments with the individual patient (Ali and Aittokallio 2019; Shi et al. 2017), rather than using monotherapy that treats all patients. In real world, treatment response is heterogeneous. Therapy should be tailored with the best response possible and highest safety margin to ensure that the right therapy is offered to “the right patient at the right time”(Subbiah and Kurzrock 2018). Precision oncology can substantially improve progress and treatment outcomes of patients. It plays a central role in revolutionizing cancer research. Consequently, alternative to calculating the average effect of an intervention over a population, many recent methods attempt to estimate individualized treatment effects (ITEs) or conditional average treatment effects from observational data (Makar et al. 2019). To accurately estimate the individualized treatment effects and learn optimal treatment choices are a key issue for precision oncology. More accurate estimation of individualized treatment effects, which provide information to guide the individual selection of the target therapies, is essential for the success of precision medicine (Kornblau et al. 2009).

Methods for estimation of individualized treatment effects (ITEs) using observational data largely differ from standard statistical estimation methods. Estimating ITEs and learning optimal treatment strategies raise a great challenge for the following reasons. First, a common framework for treatment effect estimation is the potential outcomes assumptions (Ray and Szabo 2019) where every individual has two ‘potential outcomes’ covering the hypothesized individual’s outcomes with and without treatment. Estimation of ITEs requires estimation of both factual and counterfactual outcomes for each individual. However, only the factual outcome is actually observed. We never observe the counterfactual outcomes (Rosenbaum and Rubin 1983; Chen and Paschalidis 2018; Yoon et al. 2018).

If the effect of each treatment in the subpopulation which is separately estimated is taken as an individual effect, this can create large biases. The estimated effect of each treatment in the subpopulation is still the average effect of the treatment in that subpopulation and is not individualized treatment effect in the subpopulation.

Second, clinical data often have many missing values. Simultaneously imputing both counterfactual values and missing values is not easy. Third, the function forms of the treatment effects which are often nonlinear functions are unknown (Ray and Szabo 2019). Statistical methods and computational algorithms that can efficiently deal with unknown forms of nonlinear functions are still lacking (Lengerich et al. 2019).

Classical works such as random forest and hierarchical models are adapted to estimate heterogeneous treatment effects (Wager and Athey 2015). Recently, machine learning and neural network methods are used to move away from average treatment effect estimation to personalized estimation (Alaa et al. 2017; Johansson et al. 2016; Shalit et al. 2016). AI and causal inferences are becoming a driving force for innovation in precision oncology (Seyhan and Carini 2019). A key issue for ITE estimation is to learn unobserved (missing) counterfactuals. The idea of using generative adversarial networks (GANs) for handling missing data is a very promising approach to imputing counterfactual (Goodfellow et al. 2014; Yoon et al. 2018a). Using conditional GAN (CGAN) to estimate the individualized treatment effects (GANITE) has been developed (Yoon et al. 2018a). The CGANs consist of a generator and a discriminator. The generator (G) observes the factual part of real data and imputes the counterfactuals (missing part) conditioned on observed factual data, and outputs the complete dataset. The discriminator (D) inputs the real dataset and tries to determine which part was actually observed and which part was imputed counterfactuals. The discriminator enforces generator to learn the desired distribution (hidden data distribution) (Yoon et al. 2018b).

However, the original GANITE was designed for estimation of the effects of binary treatment and cannot be applied to continuous and categorical treatments. The treatment variable in the original GANITE is binary variable which only represents the presence and absence of treatment. Therefore, the treatment variable in the original GANITE is unable to quantify the dosage of the treatment, and hence the original GANITE cannot be applied to continuous treatment. To overcome this limitation, we introduce treatment assignment indicator variable and treatment quantity variable. The treatment quantity variable can represent binary treatment, categorical treatment and continuous treatment. We change mathematical formulations of the generator and discriminator and extend GANITE from binary treatment to all types of treatments including binary, categorical and continuous treatments. The modified GANITE is abbreviated as MGANITE.

The GANITE or in general, CGAN has not systematically investigated the estimation of ITE for chemotherapy and other type of treatments in Cancer and compare the results from causal inference using observed data with the results of randomized clinical trial. One of our goals in this manuscript is to examine whether MGANITE still works well in cancer research.

In MGANITE, biomarkers that serve as conditioned variables, will be used to estimate the ITEs of both single and multiple treatments (Yoon et al. 2018a; Mirza and Osindero 2014). Sparse techniques will be employed to select biomarkers for prediction of treatment effects and to learn optimal treatment choices of patients (Emmert-Streib F, Dehmer 2019).

In summary, Novelty of the modified GANITE (MGANITE) is summarized as below.

1. The previous conditional generative adversarial network (CGAN)-based causal inference methods (GANITE) only can estimate the individualized effects of binary treatment and cannot estimate the individualized effects of continuous treatments. The proposed MGANITE is the first time to use modified CGANs for estimation of individualized effects of continuous treatments.
2. This is the first time to apply CGANs to estimation of ITE in cancer research.
3. We use randomized clinical tried data to validate the CGAN-based methods for estimation of ITE using observational data.
4. We develop new network structures for generator and discriminator in the CGANs.
5. We combined sparse techniques for selection of biomarkers with MGANITE to predict the best treatment for each patient

To evaluate its performance for estimating ITEs, simulations are conducted to estimate ITEs using simulated data and the MGANITE, and to compare its estimation accuracy with seven other state-of-the-art methods (LR, KNN, BLR, RF, and SVM). To further evaluate its performance, the MGANITE is applied to 256 newly diagnosed acute myeloid leukemia (AML) patients, treated with high dose ara-C (HDAC), Idarubicin (IDA) and HDAC+IDA at M. D. Anderson Cancer Center to estimate ITEs and identify the optimal treatment strategy for each patient. Preliminary results from simulations and real data analysis show that the MGANITE outperforms five other state-of-the-art methods. A program for implementing the proposed MGANITE for ITE estimation and optimal treatment selection can be downloaded from our website https://sph.uth.edu/research/centers/hgc/xiong/software.htm.

## Materials and Methods

### Potential outcome framework for estimation of treatment effects

We assume the Rubin causal model for estimation of treatment effects (Rubin 1974) and modified the approach to individualized treatment effect estimation in Yoon et al (Yoon et al, 2018a). The original GANITE only can estimate ITE of binary treatments, but cannot be applied to categorical and continuous treatments. We develop MGANITE which can estimate ITE of all types of treatments including binary, categorical and continuous treatments by introducing treatment assignment indicator variable and changing formulation of generator and discriminator. Consider *K* treatments. Let *T*_*k*_ be the *k*^*th*^ treatment variable that can be binary, categorical or continuous, and *T* = [*T*_1_, …, *T*_*K*_]^*T*^ be the treatment vector. We assume that there is precisely one non-zero component of the treatment vector *T*, which is denoted by *T*_*η*_, where *η* is the index of this component. Each sample has one and only one assigned treatment *T*_*η*_. To extend the binary treatment to including categorical and continuous treatments, we define the treatment assignment indicator vector *M* = [*M*_1_, …, *M*_*k*_, …, *M*_*K*_]^*T*^ as

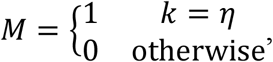

where 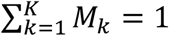.

For example, if

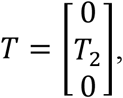

then *η* = 2 and

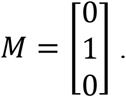

If we consider treated and untreated cases, then *K* = 2. Let *T*_1_ denote the treatment and *T*_2_ denote no treatment where *T*_2_ = 1. For the sample with the treatment, we have

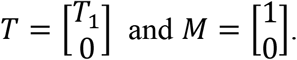

For the sample with no treatment, we have

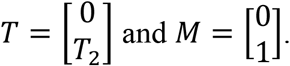

Define the vector of potential outcome *Y*(*T*) = [*Y*(*T*_1_), …, *Y*(*T*_*K*_)]^*T*^, where *Y*(*T*_*k*_) is the potential outcome of the sample under the treatment *T*_*k*_. When *K* = 2, the potential outcome *Y*(*T*_1_) corresponds to the widely used notation for one treatment *Y*^1^, the potential outcome of the treated sample, while the potential outcome *Y*(*T*_2_) corresponds to *Y*^0^, the potential outcome of the untreated sample. Only one of the potential outcomes can be observed. The observed outcome that corresponds to the potential outcome of the individual receiving the treatment *T*_*η*_ is denoted by *Y*(*T*_*η*_). The observed outcome is called the factual outcome and unobserved potential outcomes are called counterfactual outcomes, or simply counterfactuals. For the convenience of notation, the factual outcome is also denoted by *Y*_*f*_ and the counterfactuals are denoted by *Y*_*cf*_.

The observed outcome *Y*_*f*_ can be expressed as

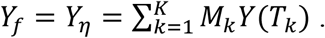

When *K* = 2, we have *M*_2_ = 1 − *M*_1_. The above equation becomes

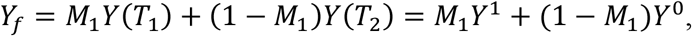

which coincides with the standard expression of the observed outcome for one treatment.

Let *X* = [*X*_1_, …, *X*_*q*_]^*T*^ be the *q*-dimensional feature vector. Assume that *n* individuals are sampled. Let 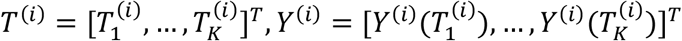 and 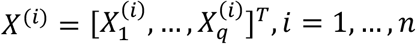 be the treatment vector, the vector of potential outcomes, and feature vector of the *i*^*th*^ individual, respectively.

The most widely used measure of the treatment effect for the multiple treatment is the pairwise treatment effect. The individual effect 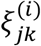 between the pairwise treatments: *T*_*j*_ and *T* _*k*_ is defined as 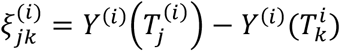, the average pairwise treatment effect 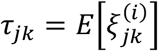. The average pairwise treatment effect *τ_j_k*|*T_j_* on the patients treated with *T*_*j*_ is defined as 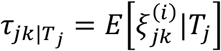

The focus of this paper is on the conditional distribution of treatment effect, given the feature vector *X*. Let *F*_*Y*|*X*_(*T*_*k*_) be the conditional distribution of the potential outcome *Y*(*T*_*k*_) under the treatment *T*_*k*_, given the feature vector *X*, and *F*_*Y*|*X*_(*T*) be the conditional joint distribution of the potential outcome vector *Y*(*T*) under the *K* treatment *T*, given the feature vector *X*. Assume that *n* individuals are sampled. For the *i*^*th*^ individual, *T*_*η*_ treatment (*M*_*η*_ = 1) is assigned. Let *X*^(*i*)^ and 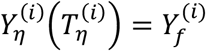 be the observed feature vector and the observed potential outcome of the *i*^*th*^ individual. Therefore, the observed dataset is given by 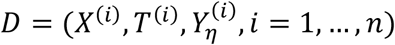. The factual and counterfactual outcomes of the*i*^*th*^ individual are denoted by 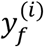 and 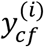, respectively.

To estimate the treatment effects, we often make the following three assumptions (Rubin 1974; Yoon et al. 2018a):

Assumption 1. (Ignorability Assumption). Conditional on *X*, the potential outcomes, *Y*(*T*) and the treatment *T* are independent,

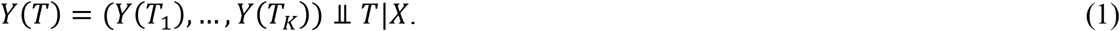

This assumption requires no unmeasured confounding variables.

Assumption 2. (Common Support). For the feature vector *X* and all treatment,

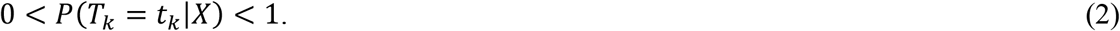

Assumption 3. (Stable Unit Treatment Value Assumption). No interference (units do not interfere with each other).

### Conditional generative adversarial networks as a general framework for estimation of individualized treatment effects

The key issue for the estimation of individualized treatment effects is unbiased counterfactual estimation. Counterfactuals will never be observed and cannot be tested by data. The true counterfactuals are unknown. Recently developed generative adversarial networks (GANs) started a revolution in deep learning (Luo and Zhu 2017). GANs are a perfect tool for missing data imputation. An incredible potential of GANs is to accurately generate the hidden (missing) data distribution given some of the features in the data. Therefore, we can use GANs to generate counterfactual outcomes.

GANs consist of two parts: “generative” part that is called the generator and “adversarial” part that is called the discriminator. Both generator and discriminator are implemented by neural networks. Typically, a *K*-dimensional noise vector is input to the generator network that converts the noise vector to a new fake data instance. Then the generated new data instance is input to the discriminator network to evaluate them for authenticity. The generator constantly learns to generate better fake data instances while the discriminator constantly obtains both real data and fake data and improves accuracy of evaluation for authenticity.

#### Architecture of conditional generative adversarial networks (CGANs) for generating potential outcomes

Features provide essential information for estimation of counterfactual outcomes. Therefore, we use conditional generative adversarial networks (CGANs) (Mirza and Osindero 2014) as a general framework for individualized treatment effect (ITE) estimation. The CGANs for ITE estimation consist of two blocks. The first imputation block is to impute the counterfactual outcomes. The second ITE block is to estimate distribution of the treatment effects using the complete dataset that is generated in the imputation block. The architecture of CGANs is shown in Figure 1.

**Figure 1.**
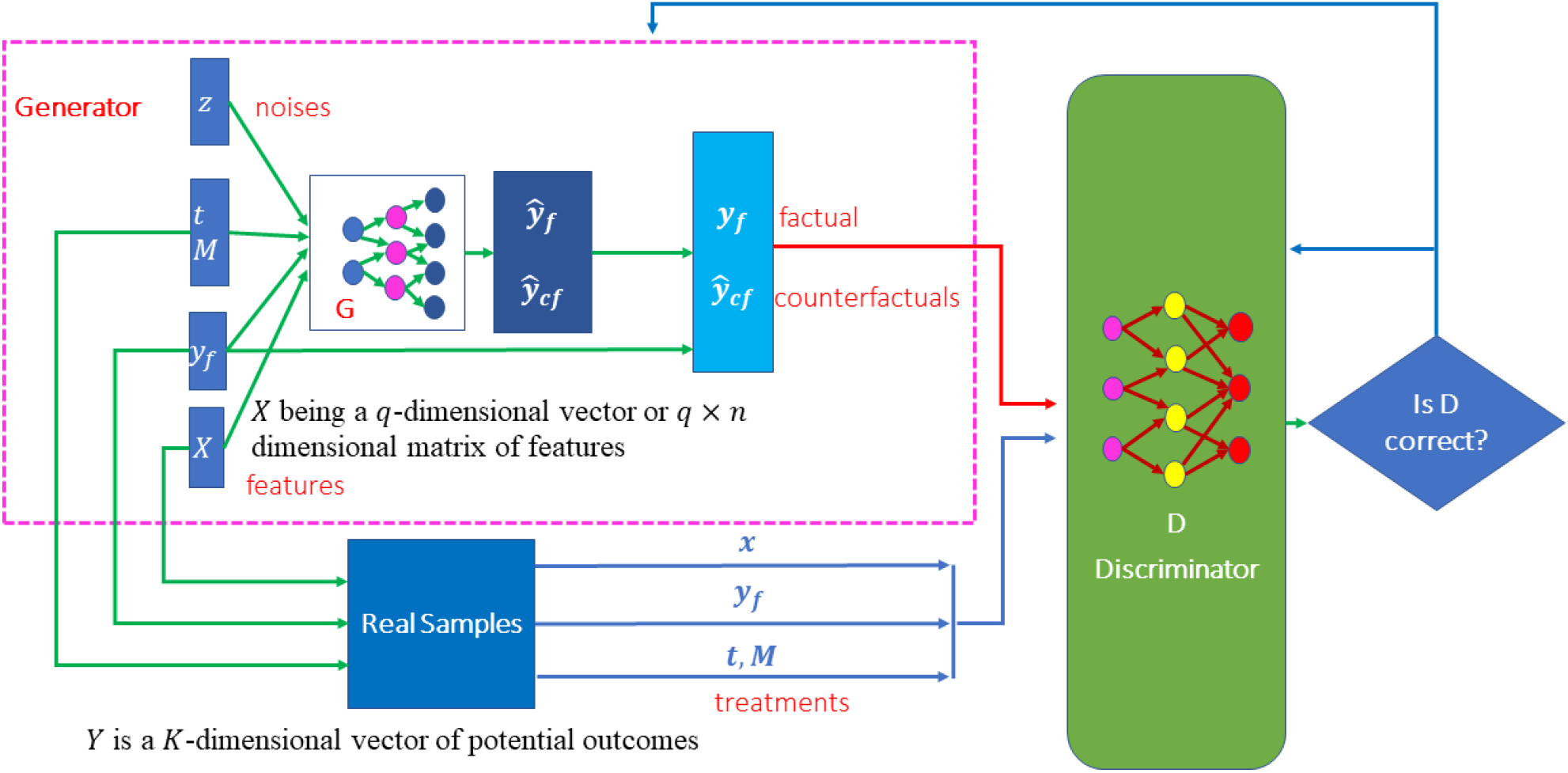
Scheme of MGANITE for the estimation of potential outcomes.

Both generator and discriminator are implemented by feedforward neural networks. The architectures of the neural networks are described as follows. The generator consists of seven layers of feedforward neural network. The first layer is the covariate input layer that input a vector × of covariates. The second and third layers are hidden layers, each layer with 64 nodes. The fourth layer concatenated the output of the third layer, the response vector Y, treatment vector T and treatment assignment indicator vector M and noise vector Z. The fifth and sixth layers are hidden layers, each layer with 64 nodes. Finally, the seventh layer is the output layer. All activation function of neurons were sigmoid function.

The architecture of the discriminator is similar to the architecture of the generator except for adding one more output layer with sigmoid nonlinear activation function.

#### Imputation block

To extend the GANTITE from binary treatments to all types of treatments, we introduce the treatment assignment vector and change some mathematic formulation of the generator. A counterfactual generator in the imputation block is a nonlinear function of the feature vector, treatment vector *T*, treatment assignment indicator vector *M*, observed factual outcome *y*_*f*_ and *K* dimensional random vector *z*_*G*_ with uniform distribution *z*_*G*_∼*U*((−1,1)^*K*^) where *Y*_*f*_ = *Y*_*η*_. The generator is denoted by

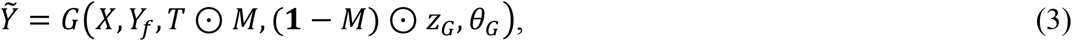

where output 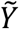 represents a sample of *G*. It can take binary values, categorical values or continuous values. **1** is a vector of 1, ⊙ denotes element-wise multiplication and *θ*_*G*_ is the parameters in the generator. We use 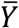 to denote the complete dataset that is obtained by replacing 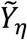with *Y*_*f*_.

The distribution of 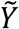 depends on the determinant of the Jacobian matrix of the transformation function *G*(*X, Y*_*f*_, *T, M, z*_*G*_, *θ*_*G*_). Changing the transformation function can change the distribution of the generated counterfactual outcomes. Let 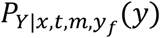 (*y*) be the conditional distribution of the potential outcomes, given *X* = *x, T* = *t, M* = *m, Y*_*f*_ = *y*_*f*_. The goal of the generator is to learn the neural network *G* such that 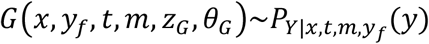.

Unlike discriminator in the standard CGANs where the discriminator evaluates the input data for their authenticity (real or fake data), the counterfactual discriminator *D*_*G*_ that maps pairs 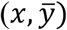 to vectors in [0,1]^*k*^ attempts to distinguish the factual component from the counterfactual components. The output of the counterfactual discriminator *D*_*G*_ is a vector of probabilities that the component represents the factual outcome. Let 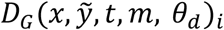 represent the probability that the *i*^*th*^component of 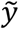 is the factual outcome, i.e., *i* = *η*, where *θ*_*d*_ denotes the parameters in the discriminator. The goal of the counterfactual discriminator is to maximize the probability 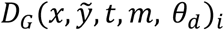 for correctly identifying the factual component *η* via changing the parameters in the discriminator neural network *D*_*G*_.

#### Loss function

The imputation block in the MGANITE attempts to impute counterfactual outcomes by extending the loss function of binary treatment in the GANITE (Yoon et al. 2018 a) to all types of treatments: binary, categorical or continuous treatments, we define loss function *V*(*D*_*G*_, *G*) as

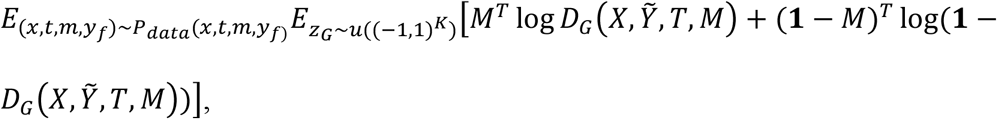

where log is an element-wise operation.

The goal of imputation block is to maximize the counterfactual discriminator *D*_*G*_ and then minimize the counterfactual generator *G*:

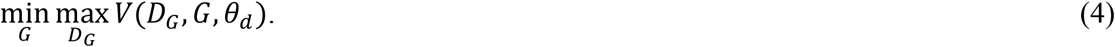

In other words, we train the counterfactual discriminator *D*_*G*_ to maximize the probability of correctly identifying the assigned treatment *M*_*η*_ and quantity of the treatment *T*_*η*_ or *Y*_*f*_(*Y*_*η*_), and then train the counterfactual generator *G* to minimize the probability of correctly identifying *M*_*η*_ and *T*_*η*_. After the imputation block is performed, the counterfactual generator *G* produces the complete dataset 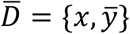. Next, we use the imputed complete dataset 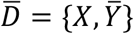 to generate distribution of potential outcomes and to estimate the ITE via CGANs which is called ITE block.

#### ITE block

The CGANs consist of three parts: generator, discriminator and loss function which are summarized as follows (Yoon et al. 2018a).

##### ITE generator

Unlike the ITE in the GANITE where the ITE generator is a nonlinear transform function of only *X* and *Z*_*I*_, the ITE generator *G*_*I*_ in the MGANITE is a nonlinear transform function of *X, T* and *Z*_*I*_:

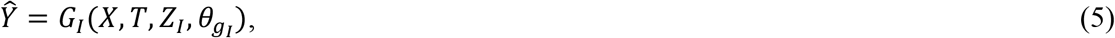

where 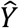 is the generated *K*-dimensional vector of potential outcomes, *X* is a feature vector, *T* is a treatment vector, and *Z*_*I*_ is a *K*-dimensional vector of random variables and follow the uniform distribution *Z*_*I*_∼𝒰((−1,1)^*K*^). The ITE generator attempts to find the transformation 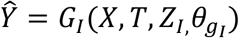 such that 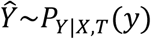.

##### ITE discriminator

Following the CGANs, we define a discriminator *D*_*I*_ as a nonlinear classifier with 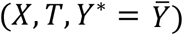 or (*X,T,Y* = Ŷ*) as input and a scalar that outputs the probability of *Y*^*^ being from the complete dataset 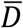.

#### Loss function

Again, unlike the loss function in the GANITE where decision function is *D*_*I*_(*X, Y*^*^), a decision function in the MGANITE is defined as *D*(*X, T, Y*^*^). The loss function for the ITE block in the MGANITE is then defined as

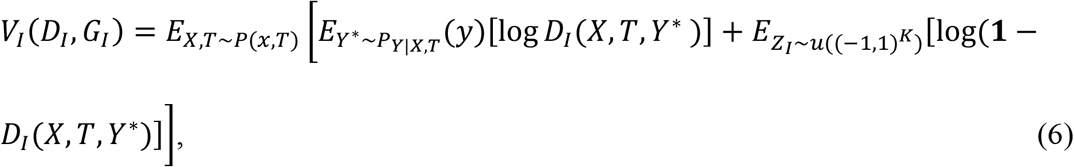

where *D*_*I*_(*X, T, Y*^*^) is nonlinear classifier that determine whether *Y*^*^ is from the complete dataset 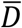 or from generator *G*_*I*_.

The goal of ITE block is to maximize the probability of correctly identifying that *Y*^*^ is from the complete dataset 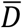 and to minimize the probability of a correct classification. Mathematically, ITE attempts

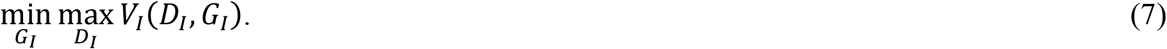

The algorithms for numerically solving the optimization problems (4) and (7) are summarized in Supplementary Note.

The learning parameters for the feedforward neural networks are given below. We set batch size equal to 16. We assumed that the learning rates for discriminator and generator were 0.0001 and 0.001, respectively. We further assume that decay rate was 0.1. Learning rate decayed (exponentially) to 10% of the starting learning rate during 70% of total batches, and stay 10% during last 30% batches. The total number of batches was 1,000,000. Adam Optimizer was used to perform optimization. We assume that 20% of the nodes are dropped randomly during training process.

### Sparse techniques for biomarker identification

The LASSO (least absolute shrinkage and selection operator) that performs both variable selection and regularization in order to enhance the prediction accuracy and interpretability of the results can be used to select biomarkers for optimal treatment selection (Ali et al. 2019). Let 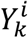 and *X*^(*i*)^denote the estimated effect of the *k*^*th*^ treatment and feature vector of the *i*^*th*^ individual, respectively. Let

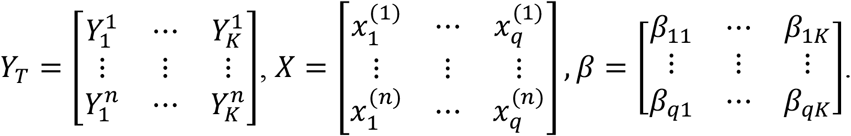

The outputs of the neural networks are in general continuous function even if the potential outcomes are binary. For the convenience of presentation, we assume that the treatment effects are continuous regardless of the potential outcomes are binary, categorical or continuous.

The LASSO estimators for identifying biomarkers that predict treatment effects are given by

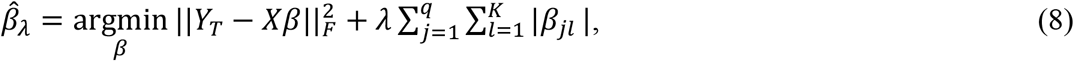

Where ‖. ‖_*F*_ is the Frobenius norm of the matrix

Nonzero elements *β*_*jl*_ ≠ 0 predict treatment effect variation and hence its correspondence *X*_*j*_ = can be used as biomarkers for investigation of the *l*^*th*^ treatment.

For the continuous treatment, we define the treatment matrix *T* and its associated coefficient matrix Γ:

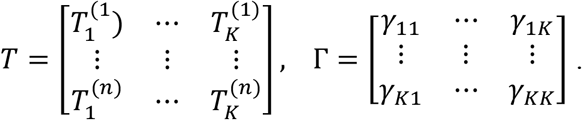

Equation (8) should be changed to

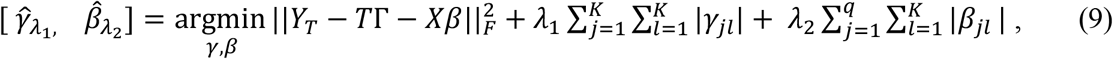

where *λ*_1_, *λ*_2_ are penalty parameters.

### Biomarker identification for optimal treatment selection

Consider *K* treatments. Let 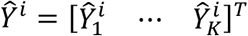 be the *K*-diemnsional vector of the estimated potential outcomes for the *i*^*th*^ individual and 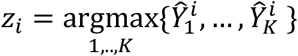 be the index of the optimal potential outcomes of the *i*^*th*^ individual. To select biomarkers for optimal treatment selection, we define the following LASSO:

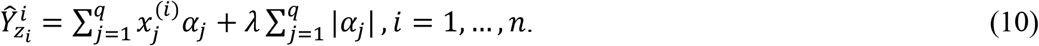

Solving the above categorical LASSO problem, we obtain a set of non-zero coefficients that are denoted as 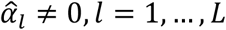, *L*. The covariates that correspond to the non-zero coefficients of the LASSO solution are chosen as biomarkers for optimal treatment selection.

Again, for the continuous treatment, equation (10) needs to be changed to

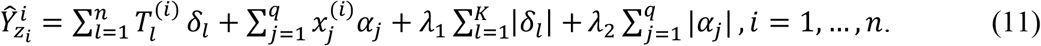

### Data collection

The proposed MGANITE was applied to 256 newly diagnosed acute myeloid leukemia (AML) patients, treated with high dose ara-C (HDAC), Idarubicin (IDA) and HDAC+IDA at M. D. Anderson Cancer Center. There were 212 valid samples and 85 useable features (14 discrete and 71 continuous), including 51 total and phosphoprotein from several biological processes such as apoptosis, cell-cycle and signal transduction pathways (Kornblau et al. 2009). Among 212 valid samples, 37 were treated with HDAC, 9 were treated with IDA and 54 were treated with HDAC+IDA, and 112 were treated by other drugs.

Prediction accuracy was defined as the proportions of correctly predicted potential outcomes. False positive rate that was defined as the proportion of individuals who were wrongly classified as positively treatment response. Discriminator accuracy is defined as proportion of correctly classified real or fake samples. Replication error is defined as cross entropy 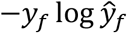 where ŷ_*f*_ = 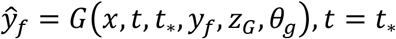, *t* = *t*_*_ and separate distance is defined as

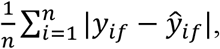

where 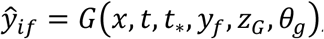, *t* ≠ *t*_*_.

## Results

### Simulations

We first examine the performance of the MGANITE in estimating the ITE of binary treatment using simulations. A synthetic dataset was generated as follows. A total of 10,000 individuals with 30-dimentinal feature vectors followed the normal distributions *N*(0, *I*). Let

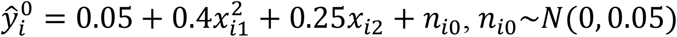

and

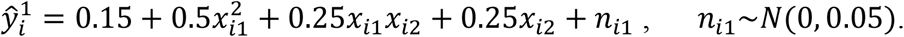

Then, the potential outcomes were generated as

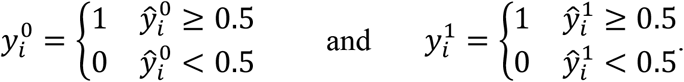

Treatment was assigned by the Bernoulli distribution:

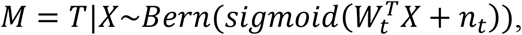

where 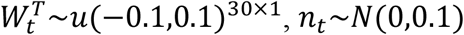, *n*_*t*_∼*N*(0,0.1), and Bern represented the Bernoulli distribution. Treatment effect can take three values of 1, 0 and -1. In other words,

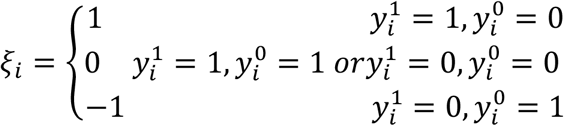

We compared MGANITE with linear regression (LR) (Makar et al. 2019), logistic regression (LogR) (Emmert-Streib and Dehmer 2019; Makar et al. 2019), support vector machine (SVM) (Makar et al. 2019), *k*-nearest neighbor (k-NN) (Crump et al. 2008), Bayesian linear regression (BLR) (Johansson et al. 2016), causal forest (CForest) (Wager and Athey 2015), and random forest (RForest) (Breiman 2001). We used six methods: MGANITE, LR, LogR, SVM, kNN and RForest to estimate the counterfactual potential outcomes and calculated the mean square error (MSE) between the estimated treatment effect and the true treatment effect, standard deviation (STD) and prediction accuracy defined as the proportions of correctly predicted potential outcomes. Table 1 presents MSE, STD and prediction accuracy of six methods to fit the generated data. We observed that the CGANs more accurately estimated the potential outcomes than the other five state-of-the-art methods. Figure 2 presented the true counterfactuals and estimated counterfactuals using MGANITE. We observed that the MGANITE reached remarkably high accuracy for estimating counterfactuals.

**Table 1.**
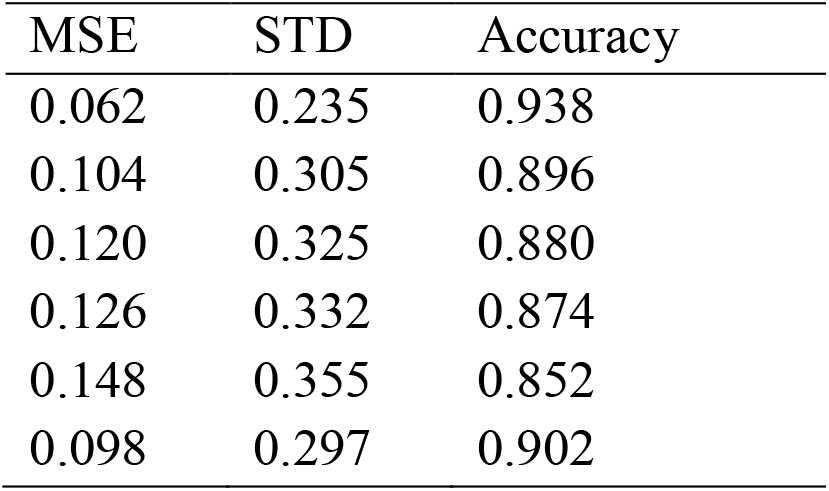
Performance of six methods for estimating the potential outcomes.

**Figure 2A.**
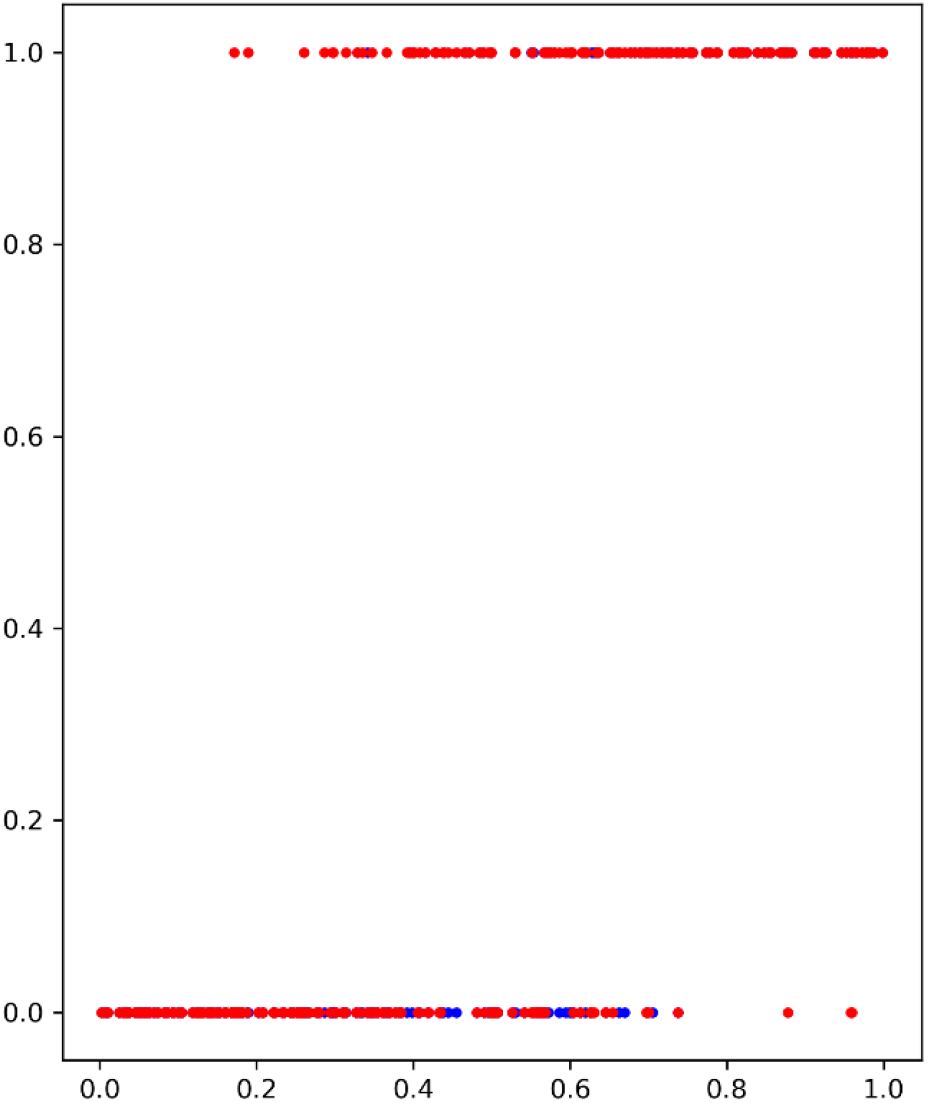
The true potential outcomes with treatment *Y*^1^ and estimated potential outcomes ŷ^1^ using MGANITE, where *x* axis denoted a value of covariate *X*_1_, *y* axis denoted the potential outcome, dot in blue color represented the true outcome *Y*^1^ and dot in red color represented the estimated outcomes ŷ^1^.

**Figure 2B.**
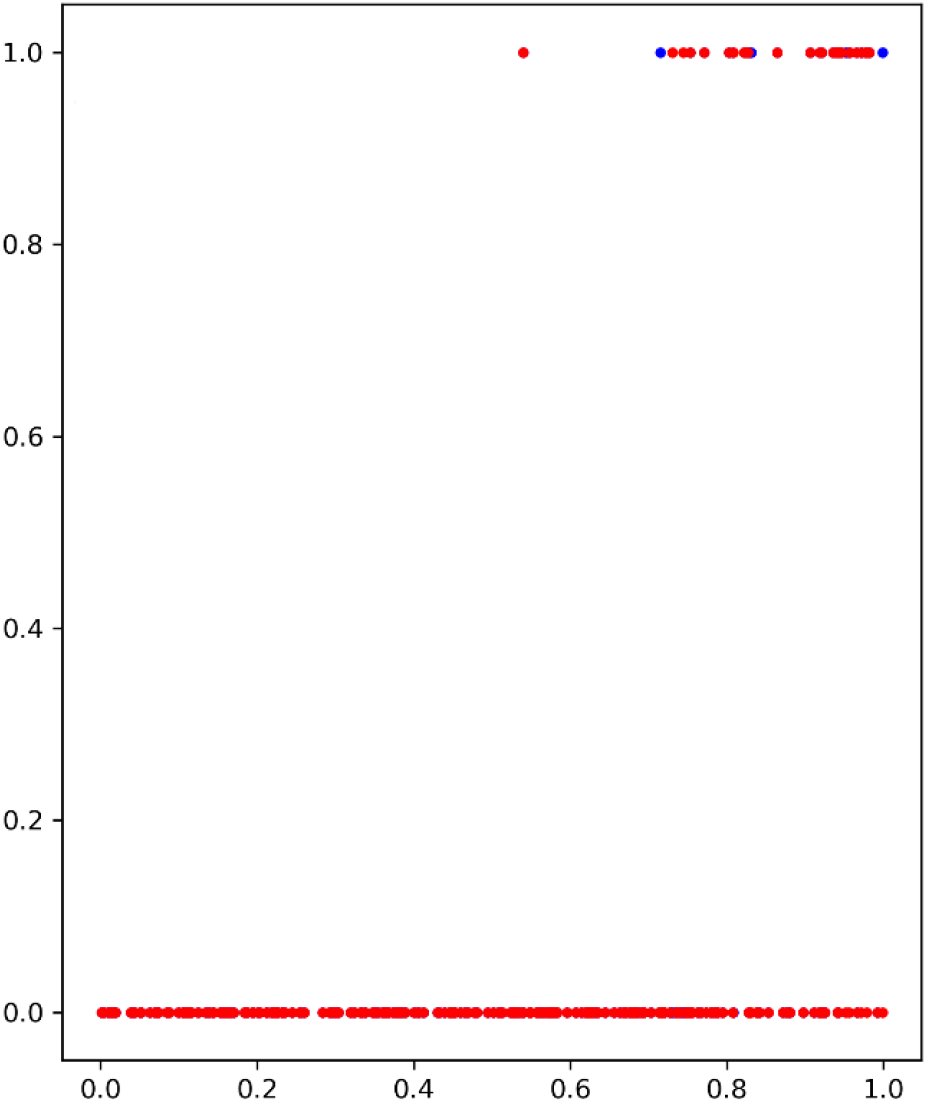
The true potential outcomes without treatment *Y*^0^and estimated potential outcomes ŷ^0^ using MGANITE, where *x* axis denoted a value of covariate *X*_1_, *y* axis denoted the potential outcome, dot in blue color represented the true outcome *Y*^0^ and dot in red color represented the estimated outcomes ŷ^0^.

The treatment effect estimation of eight methods were summarized in Table 2. Table 2 showed that the MGANITE had the highest accuracy of estimation of all treatment effects: average treatment effect (ATE), average treatment effects on the treated (ATT) and average treatment effect on the control (ATC), followed by random forests. We observed that the estimation of ATE using all methods were inflated. The inflation rates of ATE using MGANITE and RForect were 3.9% and 7.9%, respectively. The SVM reached the inflation rate of the estimation of ATE as high as 29.8%. All inflation rates of estimation of ATE using LR, LogR, SVM, KNN and BLR were very high. The simulations also showed that the false positive rate that was defined as the proportion of individuals who were wrongly classified as positively treatment response using CGANs, LR, LogR, SVC, KNN(5), KNN(10), BLR, causal forest and random forest were 3.9%, 24.7%, 28.1%, 29.8%, 28/1%, 19.7%, 25.3%, 9% and 8.4%, respectively. The results showed that false positive rates of LR, LogR, SVM, KNN and BLR for prediction of positive treatment response were too high to be applied to treatment selection. Even random forest reached the false positive rate as high as 8.4%. Table 2 also showed that the number of individuals that showed positive treatment effect increased while the number of individuals that showed no treatment effect decreased from ground truth.

**Table 2.** Treatment effects estimated for simulation data using nine methods.

| Methods | ATT | ATC | ATE | ITE=-1 | ITE=0 | ITE=1 |
| --- | --- | --- | --- | --- | --- | --- |
| Ground Truth | 0.391 | 0.321 | 0.356 | 0 | 322 | 178 |
| MGANITE | 0.399 | 0.341 | 0.37 | 0 | 315 | 185 |
| LR | 0.52 | 0.369 | 0.444 | 0 | 278 | 222 |
| LogR | 0.52 | 0.393 | 0.456 | 0 | 272 | 228 |
| SVM | 0.524 | 0.401 | 0.462 | 0 | 269 | 231 |
| KNN(5) | 0.508 | 0.401 | 0.454 | 1 | 271 | 228 |
| KNN(10) | 0.524 | 0.325 | 0.424 | 1 | 286 | 213 |
| BLR | 0.524 | 0.369 | 0.446 | 0 | 277 | 223 |
| RForest (C) | 0.452 | 0.325 | 0.388 | 0 | 306 | 194 |
| RForest (R) | 0.431 | 0.337 | 0.384 | 1 | 306 | 193 |

Next we examine the performance of the MGANITE in estimating the ITE of continuous treatment using simulations. A synthetic dataset was generated as follows.

1. Draw the covariate variable *X* from the standard normal distribution for 10,000 individuals.
2. The treatment *T* is exponentially distributed as *P*(*t*) = *e*^−(*t*−1)^, *t* ≥ 1. Define *g*(*t*) = 0.1*t*^2^.
3. Define a nonlinear function 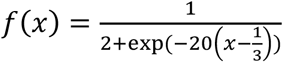
4. Define 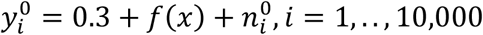, where 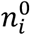 is a randomly sampled noise variable from a normal distribution *N*(0, 0.01).
5. Define 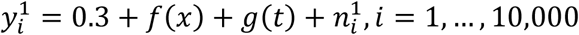, where 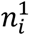 is a randomly sampled noise variable from a normal distribution *N*(0, 0.01).
6. Treatment assignment indicator variable *M*_*i*_ is drawn from a Bernoulli distribution with *P* = 0.5 for each subject.

The mean square errors (MSE) for the MGANITE, Linear Regression, KNN, Bayesian ridge regression, random forest regression and support vector machine regression were 0.011004916, 0.08500695, 0.012520364, 0.085007192, 0.014281599, 0.013962992, respectively. Figures 3 A and 3B plotted the true ITE and estimated ITE for in-samples and out-of-samples data, using six methods: MGANITE, LR, KNN, BLR, RF (R), and SVM, respectively, where dash straight line indicated that the true ITE and estimated ITE were equal. We observed from Figures 3A and 3B that many green cross points for both in-sample and out-of-sample data were much closer to the dash straight line than other types of points. This showed that the estimated ITE points using MGANITE were much closer to the true ITE point than using other five methods. In other words, the estimator of ITE using MGANITE was more accurate than that using other five methods. The results clearly demonstrated that the MGANITE outperformed 5 other state-of-the-art treatment effect estimation methods.

**Figure 3A.**
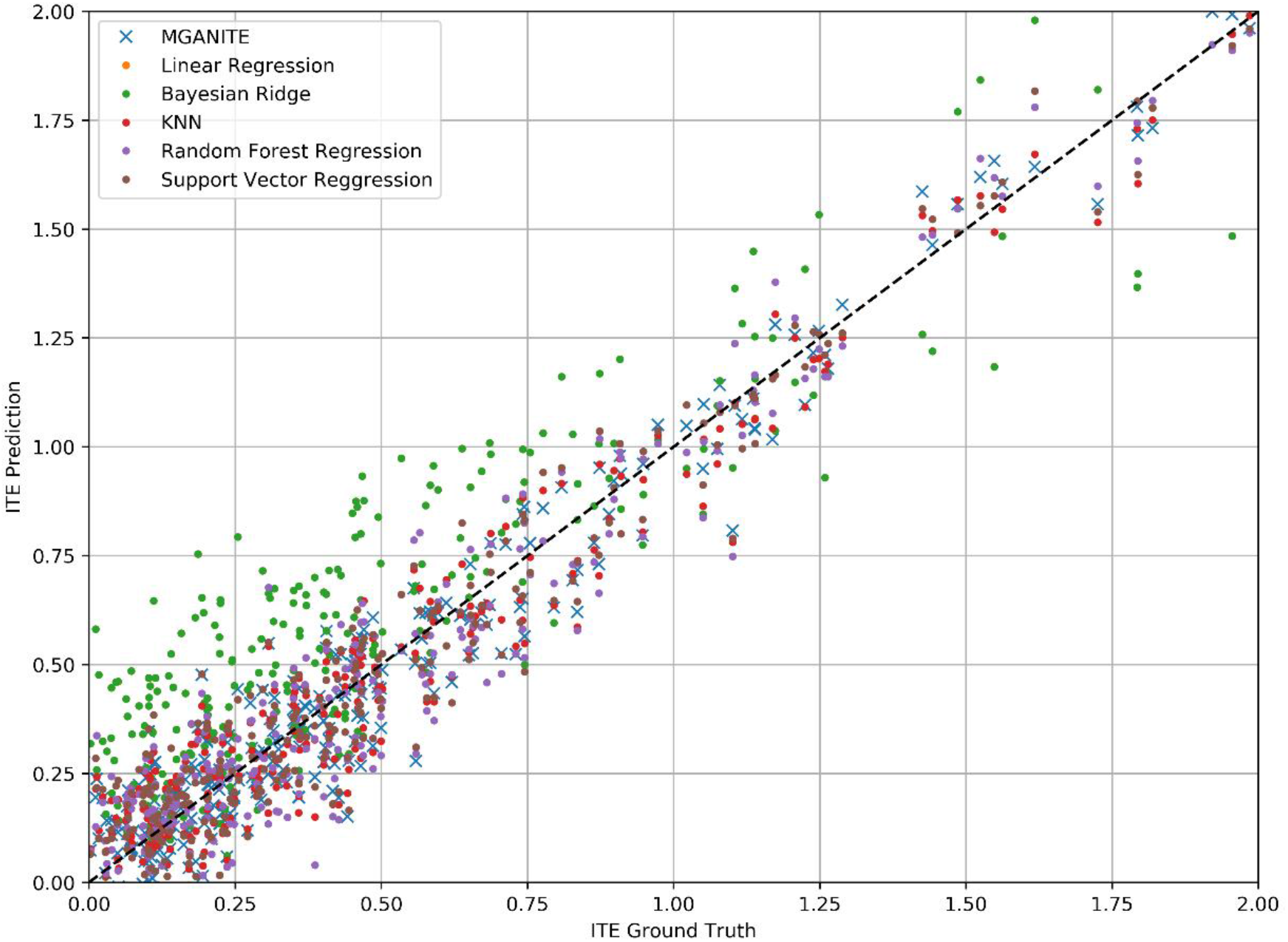
True ITE and estimated ITE for in-sample data using six methods: MGANITE, LR, KNN, BLR, RF (R), and SVM, where MGANTE was denoted by green cross point, LR was denoted by orange point, KNN was denoted by green point, BLR was denoted by red point, RF (R) was denoted by purple point and SVM was denoted by dark red point, *x* axis denoted the true ITE and *y* axis denoted the estimated ITE.

**Figure 3B.**
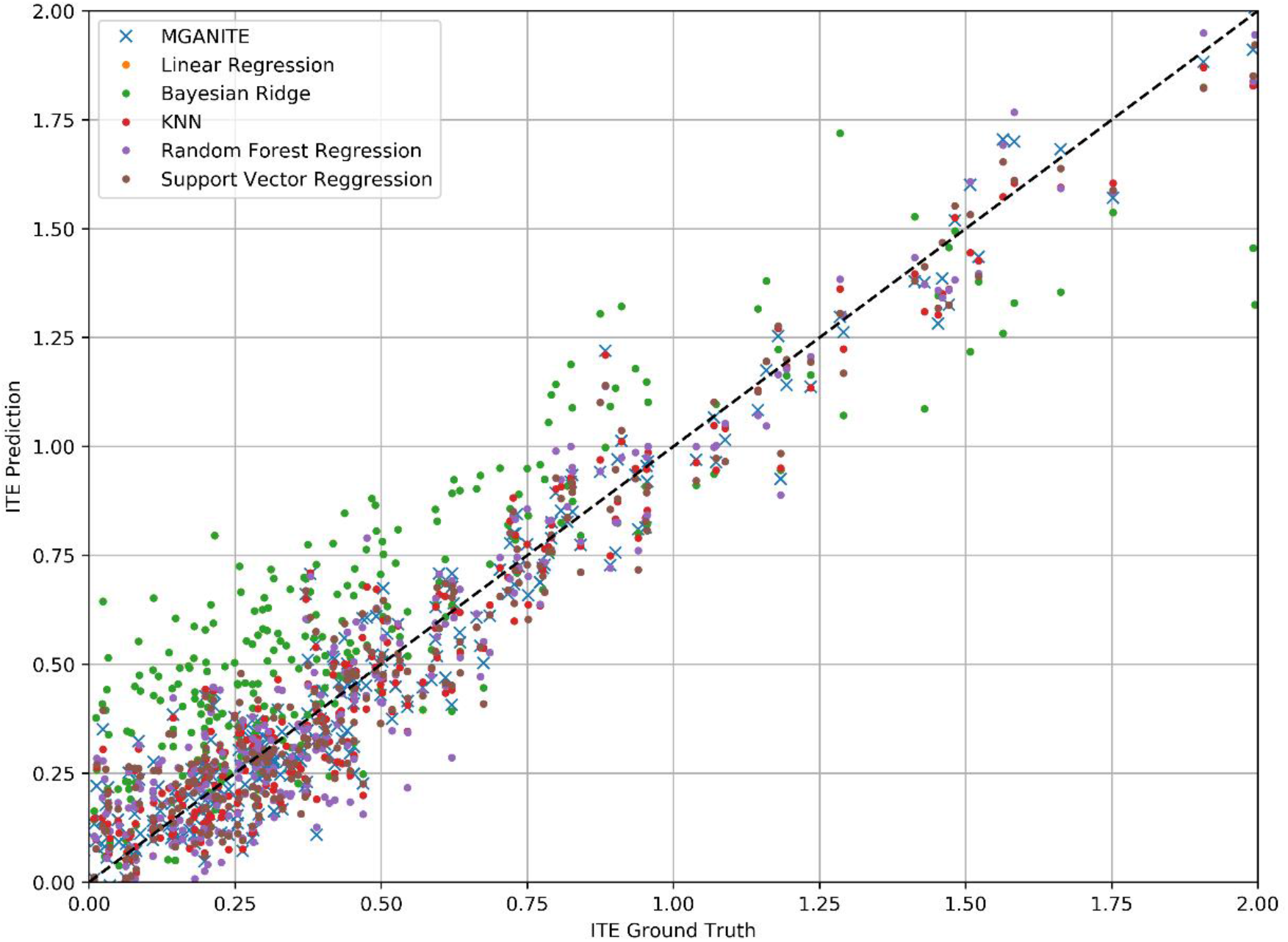
True ITE and estimated ITE for out-of-sample data using six methods: MGANITE, LR, KNN, BLR, RF (R), and SVM, where MGANTE was denoted by green cross point, LR was denoted by orange point, KNN was denoted by green point, BLR was denoted by red point, RF (R) was denoted by purple point and SVM was denoted by dark red point, *x* axis denoted the true ITE and *y* axis denoted the estimated ITE.

To further evaluate the performance of MGANITE, we provided Figure 4 that plotted receiver operating characteristic (ROC) curve for evaluation of the ability of the MGANITE to predict potential outcomes of treatment. Our calculation showed that area under the ROC curve (AUC) for MGANITE reached 0.98, a very high value. The ROC curve and AUC value demonstrated that power of the MGANITE for prediction of the potential outcomes of the treatments was very high.

**Figure 4.**
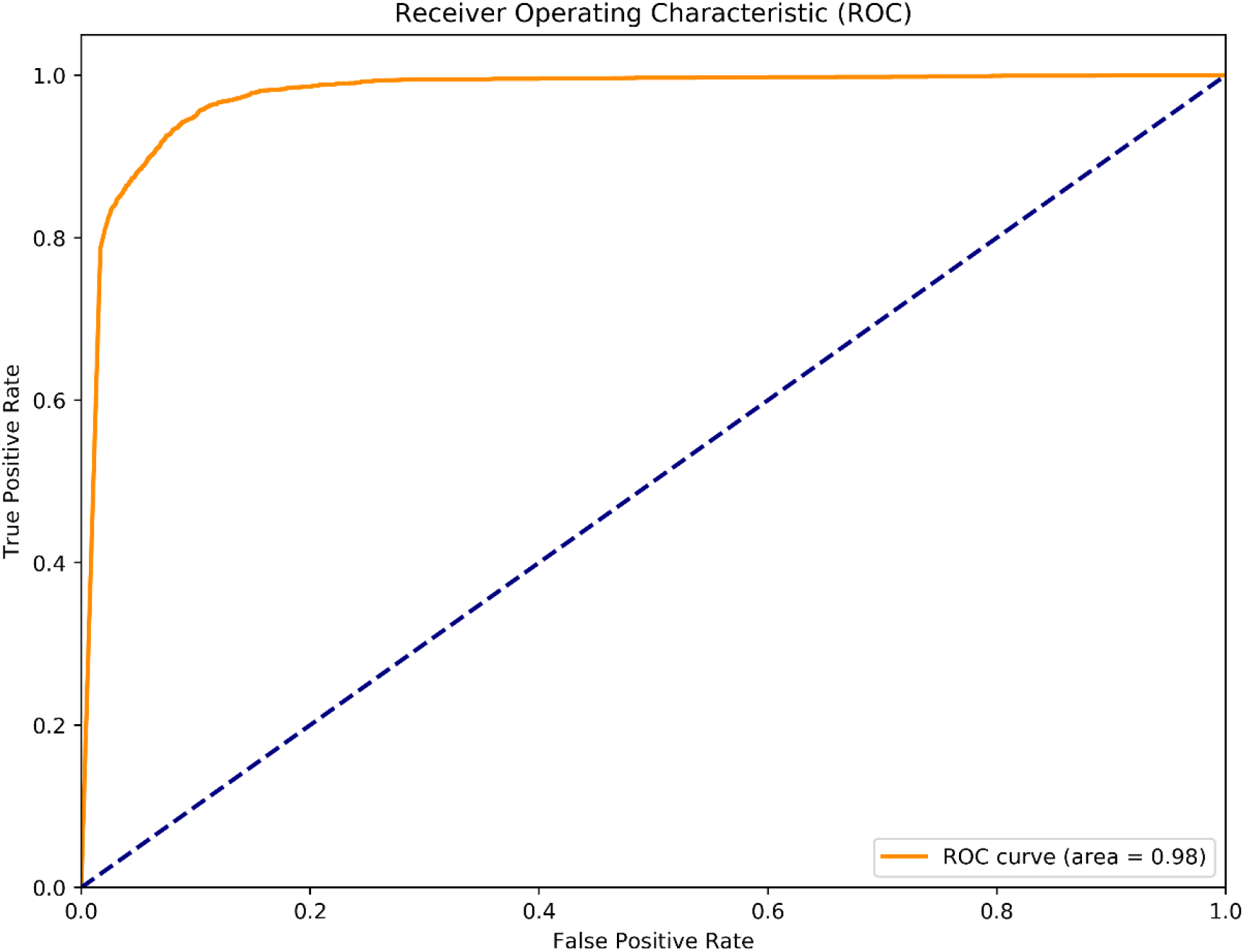
Receiver operating characteristic (ROC) curve for evaluation of performance of MGANITE.

### Real Data Analysis

The proposed MGANITE was applied to 256 newly diagnosed acute myeloid leukemia (AML) patients for clinical trial dataset (Kornblau et al. 2009).We first presented the results of treatment HDAC, HDAC+IDA (101) vs all other drugs (111). A key issue for MGANITE is how to train MGABITE. To tracking the training process of MGANITE, we presented Figure 5 that showed ATE, discriminator accuracy, replication error and separate distance curves as a function of number of batches.

**Figure 5.**
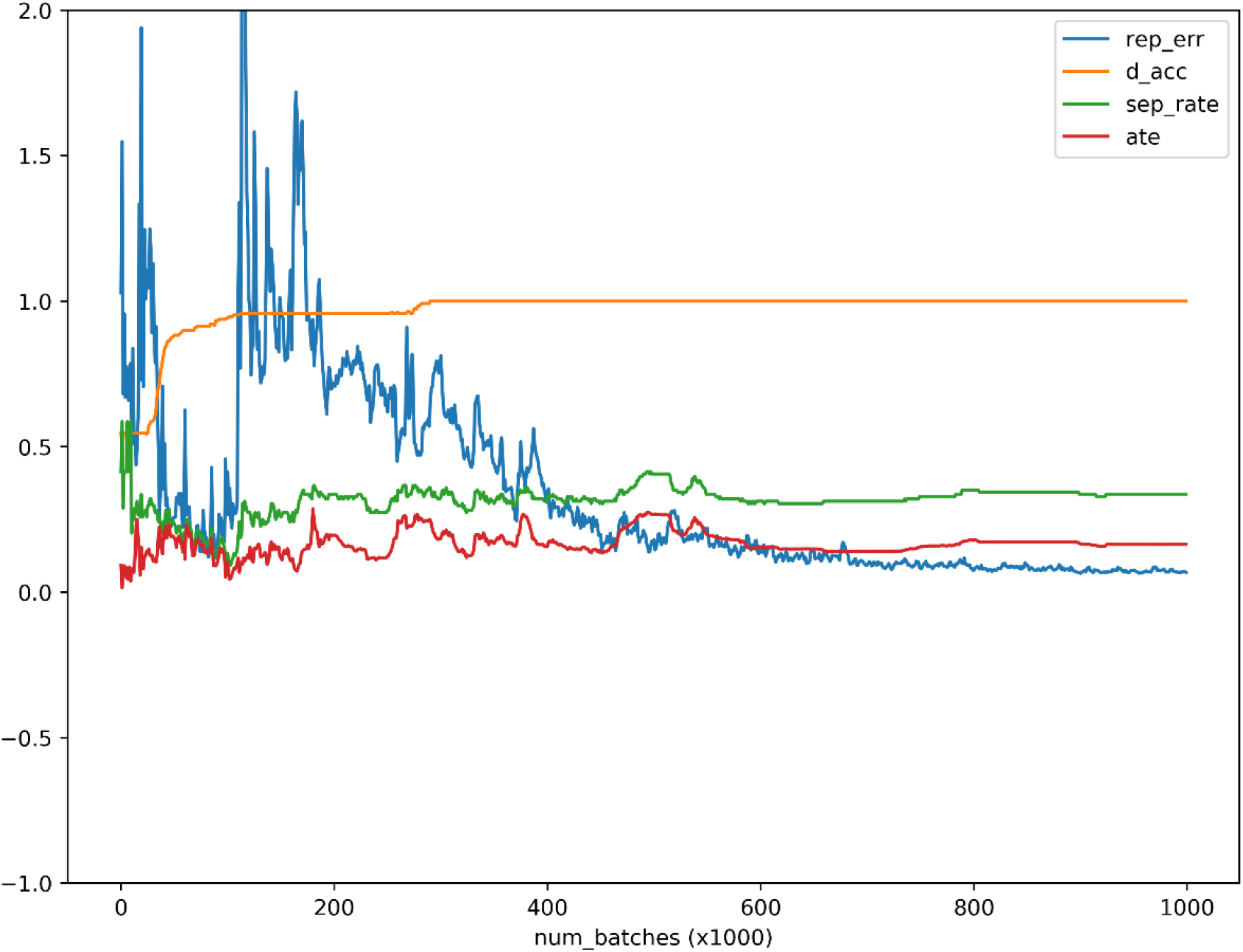
ATE, discriminator accuracy, replication error and separate distance curves as a function of number of batches where *x* axis denoted the number of batches, *y* axis denoted values of ATE, discriminator accuracy, replication error and separation distance for ATE, discriminator, replication and separation curves, respectively, red, orange, blue and green curves were ATE, discriminator, replication and separation curves.

We observed from Figure 5 that discriminator accuracy converged to 1, replication error converged to zero, separation distance converged to a constant and ATE converged to a stable value. Figure 4 demonstrated that MGANITE was trained very well.

Next we compare the treatment effect estimations using nine methods: MGANITE, LR, LogR, SVM, KNN(5), KNN(10), BLR, Rforest (C) and Rforest (R) where 5 and 10 were the number of neighbors. Treatment was HDAC or HDAC+IDA, and 85 protein expressions and other geographical variables were used as covariates. The response status (response or no response) was used as the outcome.

Table 3 summarized results of estimation of HDAC treatment effect using MGANITE and other eight methods where individuals with HDAC or HDAC+IDA were taken as the treated population, individuals with other drugs were taken as the control population. Comparison of treatment effect estimation algorithms on real data analysis is not easy because of the lack of ground truth treatment effects and small sample sizes. In general, using MGANITE, we observed that the majority of individuals who were treated by other drugs did not show any response and that 65% of the individuals who were treated by HDAC or HDAC+IDA responded. Only 13.5% of individuals who were treated by other drugs responded. To illustrate the difference between the estimated treatment effect and treatment response, we presented Figure 6 that showed the histogram of the estimated effects of the treatments HDAC or HDAC+IDA vs other drugs using MGANITE (Figure 6A), and observed the number of the response of the individuals in the population who were treated with HDAC or HDAC+IDA vs other drugs (Figure 6B). ITE was calculated based on both of factual and counterfactual. We observed that *ITE* = 0 consisted of two scenarios: (1) no response of the patients to any drugs and (2) response of the patients to both HDAC or HDAC+IDA, and other drugs. A proportion of the patients with response to HDAC or HDAC+IDA on the right side of Figure 6B and the patient with response to other drugs on the left side of the Figure 6B had *ITE* = 0. The observed response of the patients to one drug did not imply that these patients would not respond to other drugs. However, *ITE* = 1 or *ITE* = 0 implied that the patients responded to only one type of drug.

**Table 3.** Treatment effects estimated for AML dataset using nine methods.

| Methods | ATT | ATC | ATE | Number of individuals with positive treatment effect |  |  |
| --- | --- | --- | --- | --- | --- | --- |
|  |  |  |  | HDAC and<br>HDAC+IDA | No Difference | Other Drugs |
| CGANs | 0.011 | 0.356 | 0.208 | 59 | 138 | 15 |
| LR | 0.033 | 0.207 | 0.107 | 62 | 112 | 38 |
| LogR | 0.083 | 0.209 | 0.137 | 63 | 115 | 34 |
| SVM | 0.112 | 0.165 | 0.135 | 65 | 130 | 17 |
| KNN(5) | 0.248 | -0.011 | 0.137 | 55 | 131 | 26 |
| KNN(10) | 0.314 | 0.066 | 0.208 | 62 | 132 | 18 |
| BLR | 0.129 | 0.139 | 0.133 | 57 | 136 | 19 |
| RForest (C) | 0.157 | 0.286 | 0.212 | 70 | 117 | 25 |
| RForest (R) | 0.052 | 0.099 | 0.072 | 37 | 155 | 20 |

**Figure 6A.**
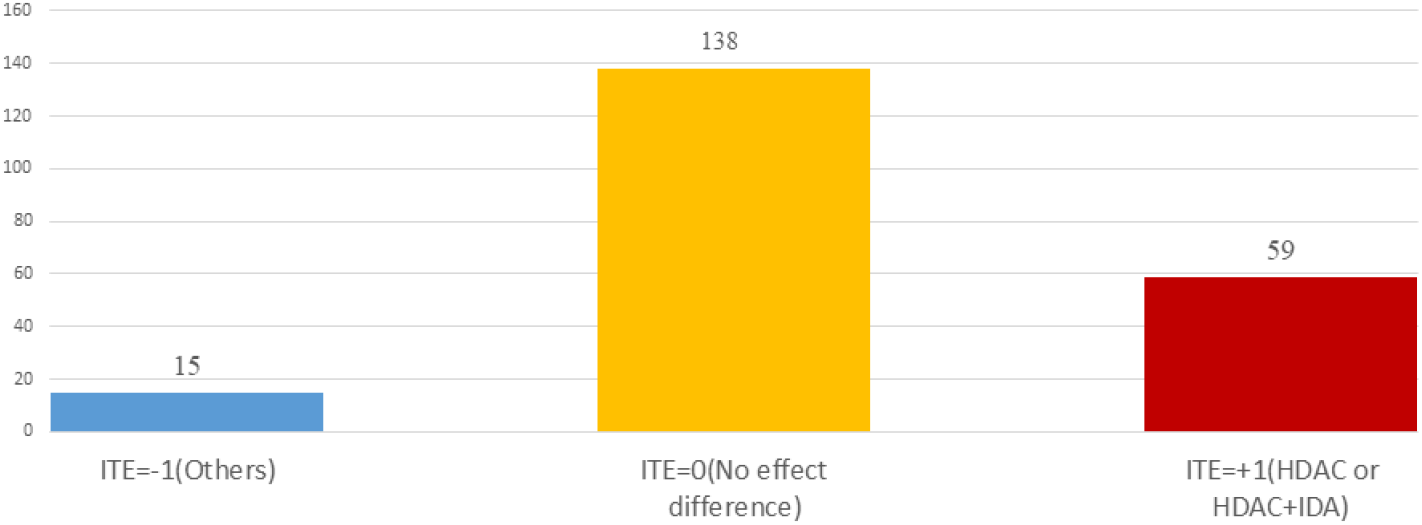
Histogram of estimated drug treatment effect using CGANs, where *x* axis denoted the value of ITE and *y* axis denoted the number of patients, *ITE* = +1 denoted the ITE of patients treated with HDAC or HDAC+IDA, *ITE* = −1 denoted the ITE of patients treated with other drugs, and *ITE* = 0 denoted the ITE of two groups of patients: one group of the patients treated with HDAC or HDAC+IDA and another group of the patients treated with other drugs.

**Figure 6B.**
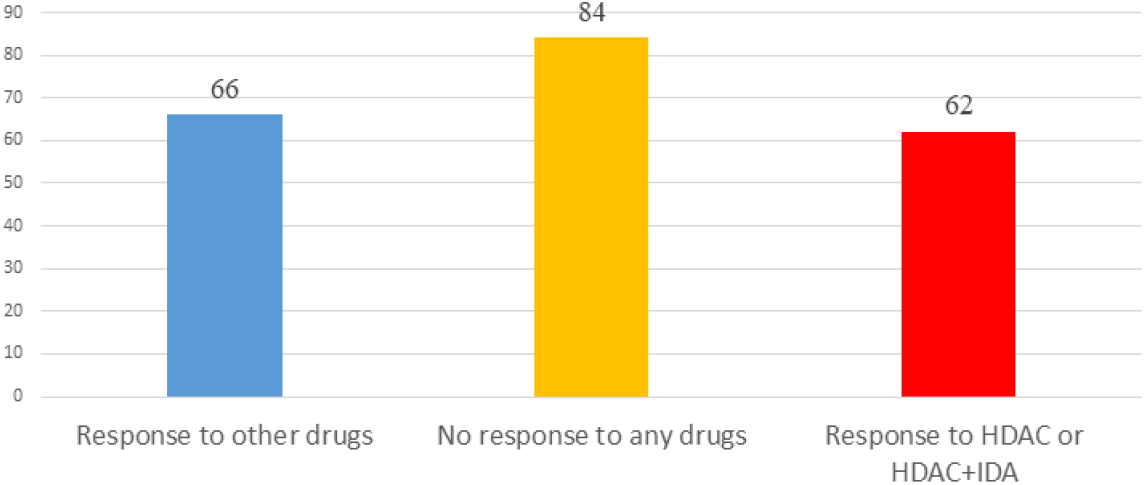
Histogram of observed drug treatment response where *x* axis indicated three scenarios as described in Figure 4B and *y* axis denoted the number of patients, the right side in the Figure 4B denoted the number of the patients only responding to the HDAC or HDAC+IDA, the middle denoted the number of the patients that responds to both (HDAC or HDAC+IDA) and other drugs or did not respond to both (HDAC or HDAC+IDA) and other drugs, and the left side denoted the number of patients only responding to the other drugs.

To further compare the performance of the MGANITE and other methods for evaluation of ITE, we split a given data set into an in-sample dataset (190 samples), used for the initial parameter estimation and model selection, and an out-of-sample dataset (22 samples), used to evaluate performance of ITE estimation. The results were summarized in Table 4. We observed that difference in the estimated ATT, ATC, ATE and proportions of ITE between In-samples and out-of-samples using MGANITE were much smaller than using other methods. This showed that the ITE estimation using MGANITE was more robust than using other methods. We calculated the Kullback-Leibler (K-L) divergence between the distributions of ITE using in-sample and out-of-samples and nine methods. The results were summarized in Table 5. Table 5 showed that K-L divergence using MGANITE was much smaller than that using other methods, which implied that MGANITE was more robust than other eight methods.

**Table 4.** Treatment effects estimated for AML dataset using nine methods.

| In-Sample |  |  |  | Proportion |  |  |
| --- | --- | --- | --- | --- | --- | --- |
| Method | ATT | ATC | ATE | ITE=-1 | ITE=0 | ITE=1 |
| MCGAN | 0.3152 | 0.2733 | 0.2911 | 0.0842 | 0.5474 | 0.3684 |
| LR | 0.1077 | -0.0210 | 0.0339 | 0.2474 | 0.4789 | 0.2737 |
| BLR | 0.0843 | 0.0817 | 0.0828 | 0.1158 | 0.6684 | 0.2158 |
| KNN(5) | -0.0247 | 0.1743 | 0.0895 | 0.1474 | 0.6158 | 0.2368 |
| KNN(10) | 0.0494 | 0.1835 | 0.1263 | 0.1211 | 0.6316 | 0.2474 |
| RF ( C ) | 0.2099 | 0.0826 | 0.1368 | 0.1421 | 0.5789 | 0.2789 |
| RF ( R ) | 0.0852 | 0.0459 | 0.0626 | 0.1316 | 0.6737 | 0.1947 |
| LogR | 0.1358 | 0.1193 | 0.1263 | 0.1579 | 0.5579 | 0.2842 |
| SVM | 0.1081 | 0.0571 | 0.0788 | 0.1158 | 0.6263 | 0.2579 |
| Out-of-Sample |  |  |  | Proportion |  |  |
| Method | ATT | ATC | ATE | ITE=-1 | ITE=0 | ITE=1 |
| MCGAN | 0.2000 | 0.3266 | 0.2691 | 0.0455 | 0.6364 | 0.3182 |
| LR | 0.4974 | 0.1222 | 0.2928 | 0.0909 | 0.5000 | 0.4091 |
| BLR | 0.3470 | 0.3129 | 0.3284 | 0.0000 | 0.5909 | 0.4091 |
| KNN(5) | 0.3000 | 0.5000 | 0.4091 | 0.0455 | 0.5000 | 0.4545 |
| KNN(10) | 0.2000 | 0.5000 | 0.3636 | 0.0000 | 0.6364 | 0.3636 |
| RF ( C ) | 0.0000 | 0.3333 | 0.1818 | 0.0455 | 0.7273 | 0.2273 |
| RF ( R ) | 0.2600 | 0.3583 | 0.3136 | 0.0000 | 0.7273 | 0.2727 |
| LogR | 0.6000 | 0.4167 | 0.5000 | 0.0000 | 0.5000 | 0.5000 |
| SVM | 0.3502 | 0.2823 | 0.3132 | 0.0000 | 0.5455 | 0.4545 |

**Table 5.** K-L divergence between the distribution of ITEs using in-samples and out-of-samples.

| Methods | Kullback–Leibler divergence |
| --- | --- |
| MGANITE | 0.00920 |
| LR | 0.04123 |
| BLR | 0.08201 |
| KNN(5) | 0.06024 |
| KNN(10) | 0.06293 |
| RF ( C ) | 0.02932 |
| RF ( R ) | 0.06407 |
| LogR | 0.09887 |
| SVM | 0.07913 |

LASSO was used to identify biomarkers for prediction of treatment effect and treatment selection. Table 6 listed the top 30 biomarkers identified by LASSO. All top 30 biomarkers explained 36.82% of variation of HDAC or HDAC+IDA treatment effect. The top Gene *GSK3* accounted for 4.4% of the explanation of treatment effect variation.

**Table 6.** Top ranking variables for explanation of treatment effect variation.

| Gene Name | R-Square (Single) | R-Square (Accumulated) | Gene Name | R-Square (Single) | R-Square (Accumulated) |
| --- | --- | --- | --- | --- | --- |
| GSK3 | 0.0440 | 0.0440 | CD33 | 0.0134 | 0.1984 |
| BILIRUBIN | 0.0411 | 0.0790 | TP53 | 0.0118 | 0.2376 |
| DIABLO | 0.0370 | 0.1266 | STAT3 | 0.0085 | 0.2415 |
| SRC | 0.0333 | 0.1329 | BIRC5 | 0.0071 | 0.2421 |
| MEK | 0.0282 | 0.1373 | BAX | 0.0070 | 0.2446 |
| AKT.p308 | 0.0244 | 0.1405 | DJI | 0.0061 | 0.2591 |
| Age_at_Dx | 0.0226 | 0.1488 | CREATININE | 0.0057 | 0.2627 |
| PRIOR_XRT | 0.0202 | 0.1776 | BAD | 0.0052 | 0.2646 |
| PSMC4 | 0.0196 | 0.1844 | ACTB | 0.0052 | 0.2816 |
| PB_Blast | 0.0181 | 0.1858 | WBC | 0.0045 | 0.2922 |
| BM_Blast | 0.0167 | 0.1878 | PRIOR_MAL | 0.0042 | 0.3190 |
| CD20 | 0.0167 | 0.1883 | FIBRINOGEN | 0.0038 | 0.3213 |
| NRP1 | 0.0147 | 0.1914 | STAT6 | 0.0033 | 0.3383 |
| TP38.p | 0.0143 | 0.1954 | CD13 | 0.0033 | 0.3409 |
| PSMC4 | 0.0135 | 0.1971 | PTEN | 0.0030 | 0.3682 |

Garson’s algorithm (Garson 1991; Zhang et al. 2018; Siu 2017) that describes the relative magnitude of the importance of input variables (biomarkers) in its connection with outcome variables (ITE) of the neural network can also be used to identify biomarkers for predicting the ITE. Top 30 biomarkers identified by Garson algorithm were listed in Table S1 where relative contribution of each biomarker to the ITE variation and cumulative contribution of biomarkers to the ITE variation were also listed in Table S1. The correlation coefficient between the importance ranking of the markers using Garson algorithm and LASSO was only -0.05.

Next, we study the joint estimation of effects of the multiple treatments. The number of individuals that were treated with HDAC, HDAC+IDA and other dugs was 37, 54 and 121, respectively. The widely used treatment estimation methods with multiple treatments are simultaneous estimation of effects of pairwise treatments. We estimated the effects of the pairwise treatments HDAC versus HDAC+IDA, HDAC versus other drugs, and HDAC+IDA versus other drugs. The results were summarized in Table 7. Pairwise comparisons listed in Table 7 did not present the results of the treatment compared with a placebo (without using any drugs). We compared the effect of one treatment with another treatment. Specifically, we made pairwise comparisons: HDAC vs other drugs, HDAC+IDA vs other drugs, and HDAC+IDA vs HDAC. The average treatment effects (ATE) of these three pairwise treatments: HDAC vs other drugs, HDAC+IDA vs other drugs, and HDAC+IDA vs HDAC using MGANITE, were 0.1001, 0.2311 and 0.1310, respectively. This demonstrated that on the average, the effect of the HDAC+IDA was the largest among the three treatments: HDAC+IDA, HDAC and other drugs, followed by the treatment HDAC. In other words, the treatment HDAC was better than other drugs, in turn, the combination of HDAC and IDA was better than HDAC. It was also noted that

**Table 7.** Multiple treatment effects estimated for AML dataset using nine methods.

|  | ATE | Number of individuals with treatment effect |  |  |
| --- | --- | --- | --- | --- |
| Method | HDAC vs Other | HDAC | No Difference | Other |
| GANITE | 0.10011 | 59 | 115 | 38 |
| LR | 0.11488 | 58 | 122 | 32 |
| LogR | 0.08962 | 54 | 123 | 35 |
| SVM | 0.14627 | 59 | 140 | 13 |
| KNN(5) | 0.18868 | 62 | 128 | 22 |
| KNN(10) | 0.35377 | 80 | 127 | 5 |
| BLR | 0.08599 | 45 | 138 | 29 |
| Rforest (C) | 0.22642 | 73 | 114 | 25 |
| Rforest (R) | 0.21274 | 58 | 145 | 9 |
| Method | HDAC+IDA vs Other | HDAC+IDA | No Difference | Other |
| GANITE | 0.23111 | 79 | 103 | 30 |
| LR | 0.0965 | 59 | 115 | 38 |
| LogR | 0.21226 | 56 | 115 | 41 |
| SVM | 0.24528 | 52 | 138 | 22 |
| KNN(5) | 0.10115 | 62 | 133 | 17 |
| KNN(10) | 0.16038 | 63 | 138 | 11 |
| BLR | 0.13066 | 49 | 137 | 26 |
| Rforest (C) | 0.07075 | 70 | 106 | 36 |
| Rforest (R) | 0.08349 | 43 | 155 | 14 |
| Method | HDAC+IDA vs HDAC | HDAC+IDA | No Difference | HDAC |
| GANITE | 0.131 | 52 | 136 | 24 |
| LR | -0.01838 | 36 | 130 | 46 |
| LogR | -0.01887 | 45 | 118 | 49 |
| SVM | -0.06278 | 9 | 181 | 22 |
| KNN(5) | 0.02358 | 34 | 149 | 29 |
| KNN(10) | -0.10849 | 11 | 167 | 34 |
| BLR | 0.01516 | 40 | 139 | 33 |
| Rforest (C) | -0.06604 | 31 | 136 | 45 |
| Rforest (R) | -0.08208 | 8 | 184 | 20 |

The effect of HDAC+IDA vs other drugs – effect of HDAC vs other drugs =0.2311-0.1001 = 0.1310 = effect of HDAC+IDA vs HDAC.

However, using LR, LogR, SVM, Rforest (C) and Rforest (R), we observed that HDAC was the best treatment. This conclusion violated the biological interpretation. We explain the reasons that caused this incorrect conclusion as follows. The traditional methods for treatment estimation are mainly based on the population average of the treatment responses. The number of observed responses and no responses of the individuals treated with other drugs was 66 and 55, respectively. The average response rate of the other drugs was 0.545. The number of observed responses and no responses of the individuals treated with HDAC was 29 and 8, respectively. The average response rate of HDAC was 0.784. The number of observed response and no response of individuals treated with HDAC + IDA was 33 and 21, respectively. The average response rate of HDAC +IDA was 0.611. Therefore, estimators of ATE of treatment HDAC vs other drugs using LR, LogR, SVM, Rforest (C) and Rforest (R) were higher than the estimators of ATE of treatment HDAC + IDA. However, the individuals treated with HDAC+IDA usually did not respond to the treatment HDAC. Therefore, the number of individuals with no response should be adjusted to be 62. After adjustment, the response rate of HDAC was changed to 0.319. Therefore, after adjustment, the ATE of HDAC vs other drugs was smaller than the ATE of HDAC +IDA. Then, the estimators of the pair-wise treatments using MGANITE were consistent with the treatment responses after the data were adjusted. This example showed that these traditional methods that were designed for single treatment effect estimation should be modified when they are applied to multiple treatment effect estimation.

Enrichment analysis to top ranking variables for explanation of treatment effect variation was performed by hypergeometric test via Reactome Pathway Database (RPD) (Jassal et al. 2020) to assess whether the number of identified biomarkers associated with Reactome pathway is over-represented than expected. Original P-value from hypergeometric test was then adjusted by FDR for multiple test correction. We found top ranking biomarkers for explanation of treatment effect variation were enriched in multiple cancer related pathways (Figure 7A), including intrinsic pathway for apoptosis (R-HSA-109606, P-value=2.86×10^−14^), Signaling by Interleukins (R-HSA-449147, P-value=2.86×10^−14^), Programmed Cell Death (R-HSA-5357801, P-value=9.7×10^−11^), PIP3 activates AKT signaling (R-HSA-1257604, P-value=2.98×10^−8^), RUNX3 regulates WNT signaling (R-HSA-8951430, P-value=1.03×10^−5^), RNA Polymerase II Transcription (R-HSA-73857, P-value=9.4×10^−5^). In addition, we could find the drug target of idarubicin (TOP2A) and Cytarabine (POLB) form significant protein-protein interaction network (P<1.0×10^−16^), indicating that the predictive biomarkers worked as the direct interactive proteins of cancer drug targets (Figure 7B).

**Figure 7.**
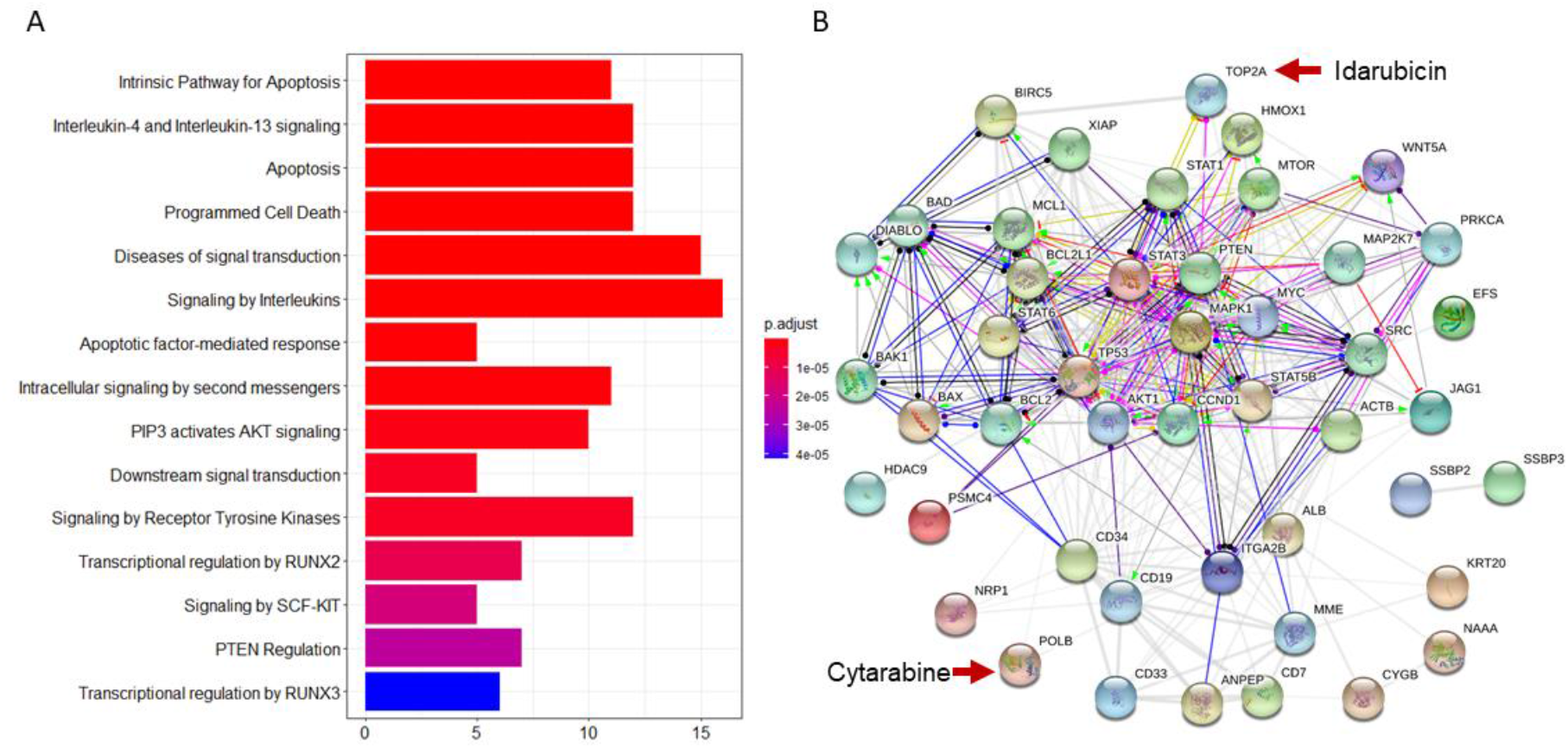
Reactome pathway analysis and protein-protein interaction (PPI) network analysis to top ranking biomarkers for explanation of treatment effect variation. **Figure 7A.** Enrichment analysis to top 44 ranking biomarkers for explanation of treatment effect variation with Reactome pathway database by hypergeometric test to assess whether the number of identified biomarkers associated with Reactome pathway was over-represented than expected. Original P-value from hypergeometric test was then adjusted by FDR for multiple test correction. Top 15 most significant enriched pathways had been showed. **Figure 7B.** PPI network analysis was performed by String 11.0 to show the protein-protein interaction among top ranking biomarkers. We found that these proteins were highly interacted which was consistent with pathway enrichment analysis (PPI enrichment P-value is 1.0e-16).

## Discussion

In this paper, we presented the MGANITE coupled with sparse techniques as a framework to estimate the ITEs and select the optimal treatments. We demonstrated that the proposed MGANITE had several remarkable features.

First, the MGANITE extended the GANITE from binary treatment to all types of treatments: binary, categorical and continuous treatments. We showed that the MGANITE had much higher accuracy for estimation of ITE than other state-of-the-art methods.

Second, in-sample and out-of-sample analysis showed that the the K-L divergence between the distributions of ITE in-sample and out-of-samples for MGANITE was much smaller than that for other methods, which implied that MGANITE was more robust than other state--of-the art methods.

Third, unlike many popular methods that are usually used to estimate the average effect of the single treatment, the MGANITE not only can estimate the ITE of single treatment, but also can accurately and jointly estimate the ITE of multiple treatments. We also showed that the results of the joint estimation of multiple treatments suing other classical methods were inconsistent and might violated the biological interpretation.

Fourth, precision oncology is the identification of the right treatment for the right patient. The essential aim is to discover biomarkers that can accurately predict individual treatment effect for each individual. Our results showed that the MGANITE with sparse techniques can identify a set of biomarkers with significant biological features. The following identified biomarkers were such typical examples.

*GSK3* is a kinase so adaptable that it has been recruited evolutionarily to phosphorylate over 100 substrates, and can regulate numerous cellular functions (Beurel et al. 2015). *GSK3* phosphorylates HDAC3 and promotes its activity, including the neurotoxic activity of HDAC3 (Bardai and D’Mello 2011). *GSK3* also phosphorylates HDAC6 to modify its activity and the link between *GSK3beta* and HDAC6 involved in neurodegenerative disorders (Chen et al. 2010).

*Bilirubin* is a reddish yellow pigment generated when the normal red blood cells break. Normal *levels* ranged from 0.2 to 1.2 mg/dL(Davis 2020). In adults, indirect hyperbilirubinemia can be due to overproduction, impaired liver uptake or abnormalities of conjugation (Gondal 2016). For AML patients, enasidenib is an inhibitor of mutant IDH2 proteins used to treat newly diagnosed mutant-IDH2 AML patients. The most common treatment-related adverse events were indirect hyperbilirubinemia (31%), nausea (23%), and fatigue (Steinwascher et al. 2015). Therefore, *bilirubin* is an important biomarker for monitoring adverse effect in AML patients who receive treatment.

Preclinical studies have discovered that Smac mimetics can directly cause cancer cell death, or make tumor cells become more sensitive to various cytotoxic treatment agents, including conventional chemotherapy, radiotherapy, or new drugs (Gulda 2015). There is synergistic interaction of Smac mimetic and HDAC inhibitors in AML cell lines, and Smac mimetic and HDAC inhibitors can trigger necroptosis when caspase activation is blocked (Meng et al. 2016).

AKT.p308 and Src.p527 are phosphorylated signal transduction proteins. These two proteins were found to have lower expression in M0, M1, M2, but they had higher levels in the other AML French-American-British (FAB) types. The expression of those two proteins, together with 22 other proteins, can be used to define distinct signatures for each FAB type (Kornblau et al. 2009).

*PTEN* is a tumor suppressor protein. Promising anti-cancer agents, HDAC inhibitors, particularly trichostatin A (TSA), can promote PTEN membrane translocation. Meng et al (Meng et al. 2016) revealed that non-selective HDAC inhibitors, such as TSA or suberoylanilide hydroxamic acid (SAHA), induced *PTEN* membrane translocation through *PTEN* acetylation at K163 by inhibiting HDAC67. Similarly, treatment with an HDAC6 inhibitor alone promoted *PTEN* membrane translocation and correspondingly dephosphorylated AKT. The combination of celecoxib and an HDAC6 inhibitor synergistically increased *PTEN* membrane translocation and inactivated AKT (Zhang and Gan 2017).

Our results showed that multiple treatments improved efficiency of drugs for curing ANL. This can be biologically explained. HDAC inhibitors have emerged as a potent and promising strategy for the treatment of leukemia via inducing differentiation and apoptosis in tumor cells (Jin et al. 2016). A phase II study with 37 refractory acute myelogenous leukemia (AML) patients showed only minimal activity of Vorinostat(HDACi), and Vorinostat failed to control the leucokyte count among most AML patients (Schaefer et al. 2009). A preclinical study revealed that the combination regimen of chidamide (a novel orally active HDAC inhibitor) and IDA could rapidly diminish tumor burden in patients with refractory or relapsed AML (Li et al. 2017). A Phase II trial of Vorinostat with idarubicin (IDA) and Ara-C for patients with newly diagnosed AML or myelodysplastic syndrome revealed good activity with an overall response rates of 85%. No excess toxicity due to Vorinostat was observed (Garcia-Manero et al. 2012). Taken together, HDACs in combination therapy with IDA or other chemotherapeutic drugs showed encouraging clinical activity in different haematologic maligancies. This explained that the combination of HDAC and IDA was the best treatment.

Although the MGANITE showed remarkable features in ITE estimation and optimal treatment selection, the results in this paper were very preliminary. Training stable GANs was a challenging task. The training process was inherently unstable, resulting in inaccurate estimation of ITEs. In this study, we ignored unobserved confounders, unmeasured variables that affect both patients’ medical prescription and their outcome. Overlooking the presence of unobserved confounders may lead to biased results. The main purpose of this paper is to stimulate discussion about how to use AI as a powerful tool to improve the estimation of ITEs and optimal treatment selection. We hope that our results will greatly increase the confidence in using AI as a driving force to facilitate the development of precision oncology.

## Data Availability Statement

Data with total of 256 newly diagnosed acute myeloid leukemia (AML) patients, treated with high dose ara-C (HDAC), Idarubicin (IDA) and HDAC+IDA were downloaded from M. D. Anderson Cancer Center (http://bioinformatics.mdanderson.org/Supplements/Kornblau-AML-RPPA/aml-rppa.xls).

## Data Availability

Data with total of 256 newly diagnosed acute myeloid leukemia (AML) patients, treated with high dose ara-C (HDAC), Idarubicin (IDA) and HDAC+IDA were downloaded from M. D. Anderson Cancer Center.

http://bioinformatics.mdanderson.org/Supplements/Kornblau-AML-RPPA/aml-rppa.xls

## Authors’ Contributions

Conception and design: M. Xiong and W. Lin

Development of methodology: Q. Ge, M. Xiong, S. Fang

Acquisition of data: X. Huang

Analysis and interpretation of data: Q. Ge, S. Guo, Y. Liu and S. Fang.

Writing, review, and/or revision of the manuscript: M. Xiong, Q, Ge, S, Guo, S. Fang, Y. Liu, W. Lin and X. Huang.

## Conflict of interest

We have no known competing financial interests or personal relationships that could have appeared to influence the work reported in this paper.

## Acknowledgements

Dr. Wei Lin is supported by the National Key R&D Program of China (Grant no. 2018YFC0116600), the National Natural Science Foundation of China (Grant no. 11925103), and by the STCSM (Grant no. 18DZ1201000). The authors thank Sara Barton for editing the manuscript and the Texas Advanced Computing Center for computation support.

---

**Table S1.** Top 30 biomarkers identified by Garson algorithm.

| Biomarkers | Ri | Sum_Ri | Biomarkers | Ri | Sum_Ri | Biomarkers | Ri | Sum_Ri |
| --- | --- | --- | --- | --- | --- | --- | --- | --- |
| CD20 | 0.0192 | 0.0192 | ALBUMIN | 0.0126 | 0.3766 | AKT | 0.0110 | 0.7250 |
| EFS | 0.0187 | 0.0380 | INFECTION | 0.0124 | 0.3890 | GSK3.p | 0.0110 | 0.7360 |
| CD7 | 0.0168 | 0.0547 | PB_Blast | 0.0122 | 0.4258 | MTOR | 0.0109 | 0.7468 |
| SSBP3 | 0.0163 | 0.0711 | S6RP.p240.244 | 0.0121 | 0.4379 | BAD.p136 | 0.0108 | 0.7577 |
| BAD.p112 | 0.0161 | 0.0871 | SURVIVIN | 0.0121 | 0.4500 | TP38.p | 0.0108 | 0.7684 |
| CREATININE | 0.0159 | 0.1031 | HGB | 0.0120 | 0.4620 | TP53 | 0.0107 | 0.7791 |
| CD10 | 0.0158 | 0.1189 | STAT3.p727 | 0.0120 | 0.4740 | ERk2.p | 0.0106 | 0.7897 |
| BILIRUBIN | 0.0157 | 0.1346 | SEX | 0.0120 | 0.4859 | BAD.p155 | 0.0105 | 0.8002 |
| CG.group | 0.0157 | 0.1503 | Source | 0.0119 | 0.4979 | STAT5.p431 | 0.0102 | 0.8104 |
| CD13 | 0.0150 | 0.1653 | BAX | 0.0119 | 0.5097 | MCL1 | 0.0101 | 0.8205 |
| AHD | 0.0146 | 0.1799 | TP | 0.0118 | 0.5215 | S6.p235 | 0.0101 | 0.8306 |
| FAB | 0.0141 | 0.1940 | BCAT | 0.0118 | 0.5333 | MEK | 0.0101 | 0.8407 |
| RACE | 0.0140 | 0.2080 | CD34 | 0.0118 | 0.5450 | ERK2 | 0.0101 | 0.8508 |
| FIBRINOGEN | 0.0136 | 0.2216 | XIAP | 0.0117 | 0.5567 | SSBP2 | 0.0100 | 0.8608 |
| CD33 | 0.0132 | 0.2348 | TP27 | 0.0117 | 0.5684 | PTEN.p | 0.0100 | 0.8707 |
| BAD | 0.0131 | 0.2479 | MEK.p | 0.0116 | 0.5800 | PTEN | 0.0099 | 0.8807 |
| S6 | 0.0131 | 0.2609 | STAT3 | 0.0115 | 0.5915 | SRC | 0.0099 | 0.8906 |
| PKCA.p | 0.0130 | 0.2740 | ZUBROD.S | 0.0113 | 0.6028 | MYC | 0.0099 | 0.9004 |
| Age_at_Dx | 0.0129 | 0.2869 | DJI | 0.0113 | 0.6140 | SMAC | 0.0096 | 0.9101 |
| NRP1 | 0.0129 | 0.2998 | PKCA | 0.0112 | 0.6252 | ACTB | 0.0095 | 0.9195 |
| PLT | 0.0129 | 0.3127 | CCND1 | 0.0112 | 0.6364 | AKT.p308 | 0.0094 | 0.9290 |
| PRIOR_MAL | 0.0129 | 0.3256 | GSK3 | 0.0112 | 0.6476 | BCL2 | 0.0093 | 0.9382 |
| WBC | 0.0129 | 0.3385 | STAT6.p | 0.0111 | 0.6587 | D835 | 0.0092 | 0.9475 |
| CD19 | 0.0128 | 0.3513 | STAT3.p705 | 0.0111 | 0.6698 | STAT1.p | 0.0091 | 0.9566 |
| PRIOR_CHEMO | 0.0127 | 0.3640 | BM_Blast | 0.0111 | 0.6809 | ITD | 0.0090 | 0.9656 |
| LDH | 0.0123 | 0.4013 | BAK | 0.0111 | 0.6920 | P70S6K.p | 0.0089 | 0.9745 |
| AKT.p473 | 0.0123 | 0.4136 | PRIOR_XRT | 0.0110 | 0.7030 | P70S6K | 0.0087 | 0.9832 |
| PB_Blast | 0.0122 | 0.4258 | BCLXL | 0.0110 | 0.7140 | SRC.p527 | 0.0086 | 0.9919 |
|  |  |  |  |  |  | MTOR.p | 0.0081 | 1.0000 |

## Notes

### Competing Interest Statement

The authors have declared no competing interest.

### Clinical Protocols

https://pubmed.ncbi.nlm.nih.gov/18840713/

### Author Declarations

This is de-identified data. Exemption of IRB/oersight.

